# A Phase II/III, Randomized, double-blind, placebo-controlled trial to evaluate Immunogenicity and Safety of the Gam-COVID-Vac combined vector vaccine in the prophylactic Treatment for SARS-CoV-2 infection in the United Arab Emirates

**DOI:** 10.1101/2024.11.29.24318199

**Authors:** Ahmed AK. Al Hammadi, Amna H. Alzaabi, Haneen B. Choker, Ahmed A. Ibrahim, Asma Bin Ishaq, Ahmed E. Mahboub, Reem S. Al Dhaheri, Mohamed N. Alzaabi, Timothy A. Collyns, Gehad ElGhazali, Stefan Weber, Basel K. al-Ramadi

## Abstract

**Background:** Different platforms were used to develop vaccines against SARS-CoV-2 which were instrumental in bringing an end to the pandemic. Nevertheless, long term efficacy and safety profiles of the various vaccine types is still being investigated. The current trial aimed to assess the safety and immunogenicity of the Gam-COVID-Vac combined vector vaccine against SARS-CoV-2-induced coronavirus infection up to 6 months post vaccination.

**Methods:** In this randomized, double-blind, placebo-controlled trial, study participants ≥18 years of age with no prior SARS-COV-2 infection or vaccination were randomized (3:1) to receive the heterologous recombinant human adenovirus-vectored vaccines serotypes 26 and 5 or placebo. Immunogenicity was determined based on quantitative IgG antibodies (Abs) to viral S and N proteins, virus-neutralizing Abs (VNA), seroconversion rates, and S protein-specific CD4 and CD8 T cell responses.

**Results:** A total of 990 participants were randomized to the vaccine (n=74) and placebo (n=246) groups. Overall incidence of Adverse events (AEs) was 1844 (81.5%) in the vaccine and 501 in placebo (82.1%) groups, the majority being in the mild/moderate categories. Two doses of vaccine induced VNA in 100% of participants on Day 42, with geometric mean ratio (GMR) reaching maximum at 120 days with average 24.14 (p<0.001). Vaccine group showed a very significant GMR for quantitative IgG to S protein. Difference in seroconversion rates was also significant with 90.0%, 83.7% and 78.9% on days 42, 120 and 180 (p<0.001 at all three points). A significant rise in the median of S protein-specific CD4^+^ and CD8^+^ T-lymphocytes with a robust IFN-γ response was evident after 28 days compared to baseline. Long-term follow-up demonstrated persistent and significant CD8^+^ T cell and IFN-γ responses at 120 days (p=0.049 and 0.039, respectively) compared to placebo group.

**Conclusion:** Gam-COVID-Vac vaccine showed a good safety profile and induced strong and durable humoral and cellular immune responses. The viral-specific CD8^+^ T cell response appears to be more durable following vaccination than CD4^+^ T cell counterpart. Importantly, there was no evidence of any severe or persistent AEs in vaccinees suggestive of long COVID syndrome, emphasizing the safety of this heterologous prime-boost vaccine.

## INTRODUCTION

Coronavirus disease 2019 (COVID-19) is an emerging respiratory infectious disease caused by severe acute respiratory syndrome coronavirus 2 (SARS-CoV-2) that infected 776,007,137 confirmed cases of COVID-19 and caused 7,059,612 deaths globally by August 18, 2024, as reported by the WHO (1). As of 26 November 2023, a total of 13.64 billion vaccine doses have been administered worldwide (1). There are currently 382 COVID-19 candidate vaccines in development worldwide. Out of the 382 candidate vaccines, 183 vaccines are in the clinical development phase (including 50 at phase III) (2). The phase III vaccine candidates include a variety of vaccine platforms such as viral vector vaccines, mRNA vaccines, inactivated vaccines, protein-subunit, DNA vaccines, virus like particle, live attenuated viruses, bacterial antigen-spore expression vectors, and viral vector + antigen presenting cells (2). Viral vector vaccines have shown efficacy in the prevention of COVID-19 infection by 68% (95% CI) and reduction in mortality with a risk ratio of 0.25 (95% CI) (3).

### The Gam-COVID-Vac Vaccine

Recombinant replication-deficient adenovirus (rAd)-based vaccines have characteristics that render them potential candidates for the WHO target product profiles for long-term protection of individuals at high risk of COVID-19 in outbreak settings since they promote a quick onset of protective immunity (4). Moreover, the use of a 2-dose immunization regimen is believed to provide a durable and long-lasting immune response (5). Prime-boost heterologous vaccination with two different vectors enables the minimization of the attenuation effect that adenoviral vectors may have on immune responses against the vector’s components (5, 6). Therefore, based on available evidence, the heterologous prime-boost vaccination approach appears to be the most efficient approach for eliciting a potent and durable immune response that is independent of the existence of an immune response to the vector (7).

The results of an interim analysis of a phase III trial involving 22,000 volunteers and using the Gam-COVID-Vac vaccine, which was carried out at a median follow-up period of 48 days post first dose, indicated an efficacy of 91.6% (95% CI 85.6–95.2) against COVID-19 infection (8). A recent meta-analysis reported that the viral vector vaccine, Gam-COVID-Vac, was likely superior to inactivated vaccines, protein-subunit vaccines, and mRNA vaccines in preventing symptomatic COVID-19 and reducing mortality (9). Randomized clinical trials that have been published indicate that viral vector vaccinations do not appear to increase the likelihood of serious adverse events (9).

### Aim of the study

The ultimate aim of worldwide R&D is to fast-track the availability of universally safe and effective vaccines. In our study, the combined vector vaccine, Gam-COVID-Vac (Sputnik V COVID-19 vaccine), was developed based on rAd type 26 (rAd26) and rAd type 5 (rAd5), both of which carry the gene for SARS-CoV-2 full-length glycoprotein S (rAd26-S and rAd5-S). The rAd26-S and rAd5-S doses are administered intramuscularly separately with a 21-day interval. The use of two varying serotypes given 21 days apart is aimed at mitigating any existing adenovirus immunity in the population (10).The aim of this study was to evaluate the immunogenicity and safety of the Gam-COVID-Vac combined vector vaccine in the prophylactic treatment for SARS-CoV-2 infection in the United Arab Emirates (UAE) (11). The study in the UAE has a distinctly different ethnic distribution. Moreover, while the original Russian study, as published in the interim analysis of 22,000 volunteers, assessed cellular immunity only 28 days after the first dose, our study extended this evaluation to day 120 after the first dose, offering an additional 92 days of data.

## METHODS

### Study design and participants

A randomized, phase II/III, double-blind, placebo-controlled trial was conducted from December 2020 through August 2021 to evaluate the immunogenicity and safety of the Gam-COVID-Vac combined vector vaccine (Sputnik V) in prophylactic treatment for SARS-CoV-2 infection. The study included participants aged at least 18 years, with negative SARS-CoV-2 PCR and IgG and IgM tests, no infectious diseases in the 14 days before enrolment. Written informed consent was obtained from each participant before screening. Screening procedure included physical examination wherein the medical history, concomitant medication, demographic, and anthropometric data were collected; and the relevant vital functions were assessed (e.g., blood pressure, heart rate, and axillar temperature). Participants were assessed by their risk category of contracting the COVID-19 infection (high, medium, and general) and underwent laboratory testing: complete blood count and biochemistry, COVID-19 diagnostics (PCR, qualitative IgM/IgG ELISA) and urine testing for drugs, alcohol, and pregnancy (in women). Volunteers participants were considered eligible if they were deemed healthy after screening procedures, had no history of COVID-19 or prior contact with patients with COVID-19 within 14 days of participation in the study, did not receive any other vaccinations, had not undergone therapy with steroids, immunoglobulins, or any other blood-derived products within 30 days, had not consumed any immunosuppressive drugs 30 days before vaccination and had no allergy to immunobiological preparations including any vaccine component. The trial was approved by the UAE Department of Health Central Ethics Committee, acting as the Local Ethics Committee (LEC; permission number DOH/CVDC/2020/2256 dated November 11, 2020), and the UAE Ministry of Health; Regulatory Committee at Ministry of Health and Prevention (RCMOHAP) permission: No. DSS-AOM-52/AMD001 dated December 14, 2020. The trial is registered with ClinicalTrials.gov (<u>NCT04656613</u>). The study is authorized and approved to be published by Human Vaccine LLC and Russian Ministry of Healthcare FGBU N.F. Gamaleya Scientific Research Center of Epidemiology and Microbiology.

### Procedures

The vaccine comprises Component I (Dose 1) consisting of recombinant adenovirus serotype 26 particles containing the SARS-CoV-2 protein S gene and Component II (Dose 2) containing recombinant adenovirus serotype 5 particles with SARS-CoV-2 protein S gene. The vaccine was manufactured as a liquid formulation containing 1x10^11^ viral particles (vp) per 0.5 ml / dose. In this phase II/III clinical trial, participants were randomized to a 1:3 ratio, with 250 participants assigned to the placebo group and 750 assigned to the vaccine group. All investigators, participants, and study staff were masked to group assignment. All randomized participants signed informed consent. After randomization, 8 participants were excluded. After exclusion, 745 remained at the end of the study of which 744 subjects took the second dose of the vaccine while 246 subjects took the second dose as placebo.

The calculated therapeutic dose of 1.0 ± 0.5 х 10^11^ v.p. was administrated intramuscularly into the deltoid muscle of the shoulder, two times on Day 1 and Day 21 and were assessed for safety, reactogenicity and immunogenicity over the whole period of investigation (180 days). Screening procedure began after the informed consent form was signed by the participants, or legal representative, and lasted no more than 7 days before a participant was determined as eligible and included in the study. Participants underwent physical examination at screening, on vaccination day (day 1 and Day 21) and on follow up days 28, 42, 120 and 180. During the observation visits, vitals were assessed in all enrolled participants, and changes in the participants’ condition and wellbeing compared to the previous visit were recorded. In addition, Adverse events were collected and reported via the weekly and biweekly Follow Up Tele-consultation/Phone calls with the participants over all the observation period up to day 180. Blood (complete blood count, Total Antibodies to SARS-COV-2, Alanine aminotransferase (ALT), Alkaline phosphatase (ALP), Aspartate aminotransferase (AST), Albumin, Total protein (TP), Total Bilirubin, Direct Bilirubin, Prothrombin time (PT), Random Glucose Blood Level, Calcium, Potassium, Bicarbonate (Total CO₂), Chloride, Blood Urea Nitrogen (BUN), Creatinine and Alcohol Level were performed at the initial screening. All participants were also tested for hepatitis B and C, HIV and syphilis once during the screening visit. Urine analysis (pH, clarity, protein, glucose, ketones, cellular content, crystals, pregnancy) was performed three times over all the study period, at the screening, before the vaccine injection on day 1 and on day 21. The following immune parameters were assessed: Virus-neutralizing activity (VNA), Quantitative IgG antibodies to S-protein, Qualitative IgG to N-protein, CD4 and CD8 T-cell proliferation response to S-protein, and IFN-γ recall response to S-protein. Quantitative IgG antibodies to S-protein, Qualitative IgG to N-protein and Serum neutralizing activity were assessed at four different time points of the study period: prior to immunization on day 1 and on days 42, 120 and 180 after immunization. For the evaluation of cell-mediated immune responses, CD4 and CD8 T-cell proliferation and IFN-γ response to S-protein were carried out prior to immunization on day 1 and on days 28 and 120 after immunization (3 time points). Biological material was collected for SARS-CoV-2 and tested using PCR on screening visit and prior to the immunization on day 21 (2 times).

### Sample size calculation and justification

Based on published data, antibody titers in the patients who recovered from COVID-19 become GMT IgG 2974.83 in >21 days (12). Sample size estimation was calculated for a repeated measures ANOVA using the G*Power software (Version 3.1.9.2). Since the estimated effect size is 0.12, for the purpose of this study a significance level of 5% (α = 0.05), and power of 90% are used. The design will adopt an unbalanced design of the placebo and vaccinated group of a 1:3 ratio. Normalizing the number of recruited subjects by 30% for adverse events and dropouts (732*1.3) the total number of subjects that will be recruited will be 1000 (250 subjects in the placebo group and 750 in the vaccinated group). Sample sizes were selected to balance statistical power and recruitment feasibility.

### Outcomes

The primary outcome measures for immunogenicity were measurement of the change from baseline of antibody levels specific to SARS-CoV-2 glycoprotein S on days 1 (before the first vaccine dose) and on days 42, 120 and 180 after first dose of vaccination; changes in SARS-CoV-2 neutralizing antibody titers at the same time points; and determination of CD4^+^ and CD8^+^ T-cell responses and IFN-**γ** production after recall stimulation with viral S protein on day 1 (prior to vaccine administration) and days 28 and 120 following vaccination. Changes in the level of antibodies against SARS-CoV-2 N-protein were also determined. Secondary outcome measure was safety of the “Sputnik V” Vaccine. Outcome measures for safety were the number and severity of adverse events during the whole study after injection of the first dose of the vaccine.

### Statistical analysis

Statistical data processing used the SPSS version 24 software (IBM Corp. Released 2016. IBM SPSS Statistics for Windows, Version 24.0. Armonk, NY: IBM Corp.) and GraphPad Prism 10 (One-way ANOVA followed by Dunnett’s multiple comparisons test was performed using GraphPad Prism version 10.0.0 for Windows, GraphPad Software, Boston, Massachusetts USA, www.graphpad.com) software application package. The statistical analysis of immunogenicity included all the participants who have completed the study as per protocol. The safety analysis included all the participants who had received a dose of the study vaccine. No replacement of missing, unanalyzable or doubtful data has been envisaged. The geometric mean concentrations of the Quantitative IgG antibodies to S and N Protein and virus-neutralizing antibodies in trial participants were computed as the exponential of the mean of the log-transformed concentrations and its 95% confidence interval was reported. The log-transformed concentrations were compared between the two study arms using the independent Student’s t-test at 42±4, at 120±14 and at 180±14 days after the first dose. Geometric mean ratios were calculated along with their 95% confidence Interval. The percentage of participants with 4-fold or more increase in the concentrations of Quantitative IgG antibodies to S and N Protein in trial subjects on the drug administration day before injecting the first dose of the study vaccine/placebo and on 42±4 days after the first dose was computed along with their 95% confidence intervals using the Wald Method or the Clopper-Person methods if needed (when at least one observed cell count is below 5). Differences between the two study arms were compared using the Chi-squared test. Cellular immune response levels were summarized using medians 25^th^ (Q1) and 75^th^ (Q3) quartiles values. Levels of IFN-γ that were below the limit of detection were replaced by the border limit of detection to be able to compute the mathematical measures in both arms. The change in the concentrations of IFN-γ and the extent of proliferative CD4^+^ and CD8^+^ T cell responses between participants in the two study groups were analyzed using the Wilcoxon-Mann-Whitney test (or the t-test on the log-transformed endpoints) at days 28±4 days and at 120±14 days after the first dose using GraphPad Prism 10.0 (San Diego, CA, USA). Increases in those variables within each study arm were tested using the Wilcoxon signed-rank test at the specified time points. The safety analysis included all participants who had received the study vaccine/ placebo injection and for whom all available data were verified in the case report form at the time of database lock. Adverse events were coded using MedDRA, version 24.0. Adverse events were presented by group, system organ and class, and preferred term. AE severity and causality have been assessed by a physician investigator. The results of laboratory examinations, vital signs, and other safety parameters are provided descriptively.

## RESULTS

### Study Participants

A total of 2239 participants were screened, out of which 1239 (55.3%) were excluded due to eligibility criteria. The remaining 1000 (44.6%) were enrolled and randomized on a 1:3 ratio with 750 assigned to the vaccine group and 250 to placebo. The demographic characteristics of the enrolled study participants are summarized in **Table I**. It is also worth noting the wide ethnic distribution of the participants in this study, which was as follows: South-East Asia 48.7%, MENA region 36.5%, Central/South America 9%, Africa 3.7%, and Europe 2.1%. This distribution is unique to the UAE. After randomization, 8 participants were excluded (5 subjects from the vaccine arm and 3 from the placebo) for not meeting eligibility criteria. The reasons for exclusion were early withdrawal due to asthma (n=3), pregnancy (n=1), positive PCR (n=2), positive IgM/IgG for SARS COV2 (n=1), or active tuberculosis (n=1). After exclusion, 745 remained of which 744 subjects took the second dose of the vaccine. Similarly, out of the remaining 247 subjects in the placebo group, 246 took the second dose. For the immunogenicity data, 17 (2.3%) were missed from the vaccine arm versus 10 (4.0%) from the placebo arm. The main reasons for withdrawal were either loss to follow-up or not attendance of the visit. Finally, 78 subjects were included in the cell-mediated study in the vaccine group versus 22 in the placebo at baseline. Six subjects didn’t attend the test at 28 days yielding a total 94 individuals with available data while 10 more subjects were absent at 120 days yielding a final total of 84 individuals. Full details are presented in **Figure 1**.

**Figure 1:**
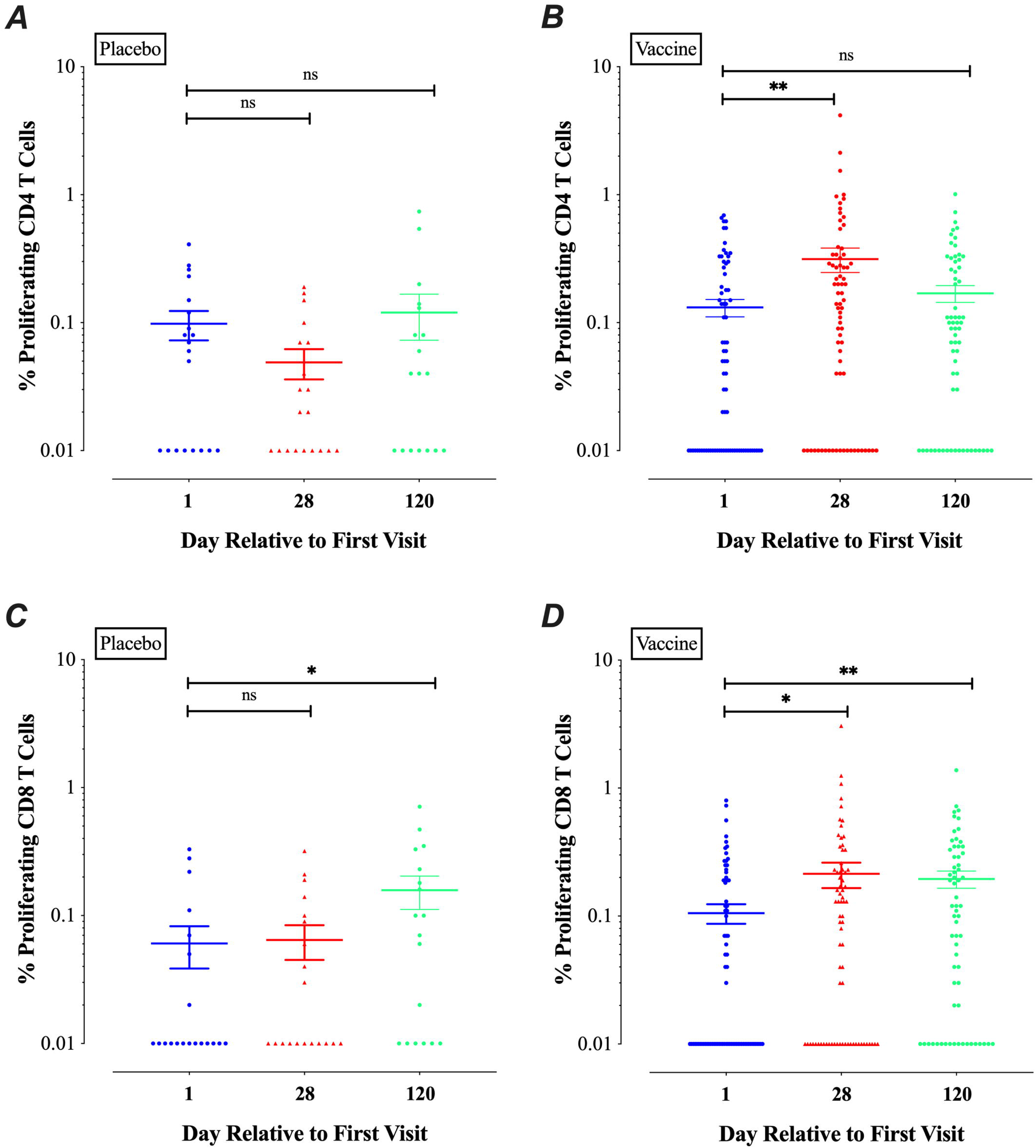
Profile of study participants and distribution

**Table I.** Demographic and anthropometric characteristics of trial participants.

| Variable | Vaccine (n=744) | Placebo (n=246) |
| --- | --- | --- |
| Sex (n, %) |  |  |
| Male | 637 (85.5) | 202 (82.1) |
| Female | 107(14.5) | 44 (17.9) |
| Age (in years) mean±SD | 34.1±8.9 | 34.9±9.9 |
| Race* (n, %) |  |  |
| Caucasian | 81 (10.9) | 28 (11.4) |
| Asian | 590 (79.3) | 196 (79.7) |
| Black | 73 (9.8) | 22 (8.9) |
| Bodyweight, kg (mean±SD) | 79.1±15.5 | 75.9±14.8 |
| Height, cm (mean±SD) | 171.1±8.6 | 170.8±8.9 |
| Body-mass index**, kg/m<sup>2</sup> (mean±SD) | 27.0±5.0 | 25.9±4.3 |
| Medical condition/Medical history (n, %) |  |  |
| Yes | 224 (30.1) | 81 (32.9) |
| No | 520 (69.9) | 165 (67.1) |
| Risk of Infection*** (n, %) |  |  |
| High | - - | - |
| Medium | 8 (1.1) | 3 (1.2) |
| General | 733 (98.7) | 243 (98.8) |

\* Race or ethnic group was reported by the participants
\*\* Calculation was based on the bodyweight and height measured at the time of screening.
\*\*\* General risk is identified that there are no additional risks related to professional activities; a medium risk is identified that there is a professional contact with many people (general medical officers, social services employees, shop assistants, etc.); while, a high risk means that the job involves interactions with the patients having confirmed COVID-19 diagnosis.

Overall incidence of AEs in the vaccine group was 1844 versus 501 in the placebo group. Most of the recorded AEs in the vaccine arm were either mild (n=460; 61.8%), or moderate (n=99; 13.3%); only 6.2% (n=46) were severe AEs. Similarly, in the placebo group, most reported AEs were either mild (n=160; 65.0%) or moderate (n=26; 10.6%), with only 6.5% (n=16) reporting severe AEs. No death was reported in either arm. Four serious adverse events were reported in the placebo arm after 42 days while five serious adverse events were reported after 120 days. At day 120 post vaccination, the majority of subjects reported mild intensity events, 307 (41.2%) in vaccine group and 90 (36.6%) in placebo group, Moderate intensity events were reported by 38 subjects (5.1%) in vaccine group and 18 subjects (7.3%) in placebo group. Few subjects reported severe intensity events, 7 (0.94%) in vaccine group and 3 (1.2%) in placebo group. Finally, one potential life-threatening event was reported by a subject from the placebo group. Details of AEs in total or following each vaccine dose are presented in Tables II, III and IV. Overall, only a few events were determined to be related to the investigational medicinal product after 120 days (44 events in vaccine group and 13 events in the placebo group). **Table V** shows more details about the relatedness of AEs to the medicinal product.

**Table II.** Adverse events and serious adverse events frequency and percent of participants.

|  | 42 days |  |  |  | 120 days |  |  |  |
| --- | --- | --- | --- | --- | --- | --- | --- | --- |
|  | Vaccine<br>N=744 |  | Placebo<br>N=246 |  | Vaccine<br>N=744 |  | Placebo<br>N=246 |  |
| **Adverse Events** | **Events #** | **No. (%) of Subjects** | **Events #** | **No. (%) of Subjects** | **Events #** | **No. (%) of Subjects** | **Events #** | **No. (%) of Subjects** |
| Any AE | 1844 | 606 (81.5%) | 501 | 202 (82.1%) | 697 | 320 (43.0%) | 268 | 100 (40.7%) |
| **AE\* intensity** |  |  |  |  |  |  |  |  |
| Mild | 1664 | 460 (61.8%) | 449 | 160 (65.0%) | 638 | 307 (41.3%) | 232 | 90 (36.6%) |
| Moderate | 119 | 99 (13.3%) | 28 | 26 (10.6%) | 52 | 38 (5.1%) | 32 | 18 (7.3%) |
| Severe | 60 | 46 (6.2%) | 20 | 16 (6.5%) | 7 | 7 (0.94%) | 3 | 3 (1.2%) |
| Potentially life threatening | 1 | 1 (0.1%) | - | - | - | - | 1 | 1 (0.4%) |
| death | - | - | - | - | - | - | - | - |
| **Any SAE\*\*** | - | - | 4 | 3(1.2%) | 3 | 3(0.4%) | 2 | 2 (0.8%) |

\*AE=Adverse Events
\*\*SAE=Severe Adverse Events

**Table III:** Frequency of adverse / serious adverse events and percent of participants after first dose.

|  | **Vaccine**<br>**N=744** |  | **Placebo**<br>**N=246** |  |
| --- | --- | --- | --- | --- |
| **Adverse Events** | **Number of**<br>**events** | **Number (%) of**<br>**subjects** | **Number of**<br>**events** | **Number (%)**<br>**of subjects** |
| Any AE* | 885 | 300 (40.3%) | 233 | 96 (39.0%) |
| AE intensity |  |  |  |  |
| Mild | 816 | 232 (31.2%) | 213 | 75 (30.5%) |
| Moderate | 48 | 48 (6.5%) | 12 | 12 (4.9%) |
| Severe | 21 | 20 (2.7%) | 8 | 8 (3.3%) |
| potentially life threatening | - | - | - | - |
| death | - | - | - | - |
| Any SAE** | - | - | 2 | 2 (0.8%) |

\*AE=Adverse Events
\*\*SAE=Severe Adverse Events

**Table IV:**
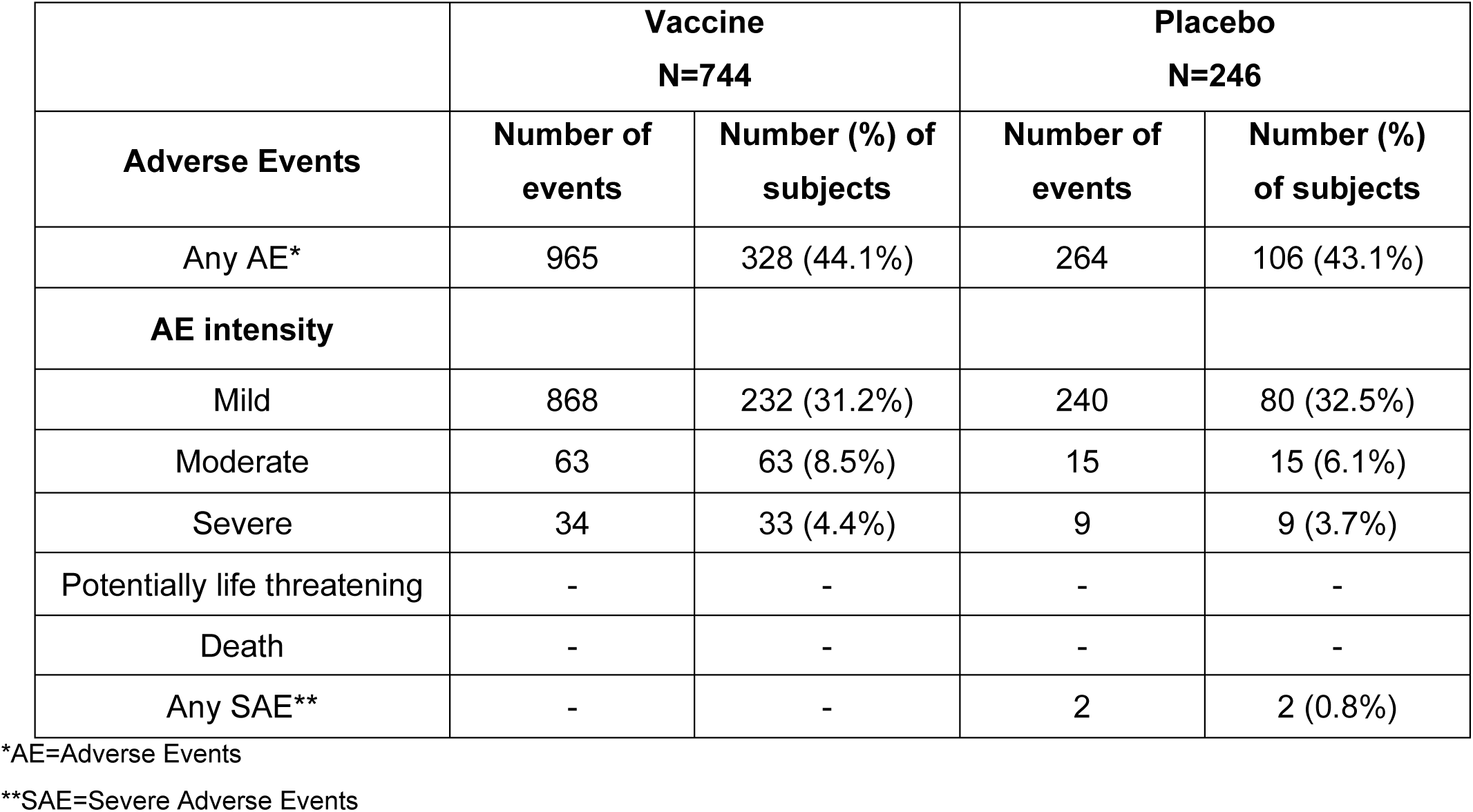
Frequency of adverse / serious adverse events and percent of participants after second dose.

|  | **Vaccine**<br>**N=744** |  | **Placebo**<br>**N=246** |  |
| --- | --- | --- | --- | --- |
| **Adverse Events** | **Number of**<br>**events** | **Number (%) of**<br>**subjects** | **Number of**<br>**events** | **Number (%)**<br>**of subjects** |
| Any AE* | 965 | 328 (44.1%) | 264 | 106 (43.1%) |
| **AE intensity** |  |  |  |  |
| Mild | 868 | 232 (31.2%) | 240 | 80 (32.5%) |
| Moderate | 63 | 63 (8.5%) | 15 | 15 (6.1%) |
| Severe | 34 | 33 (4.4%) | 9 | 9 (3.7%) |
| Potentially life threatening | - | - | - | - |
| Death | - | - | - | - |
| Any SAE** | - | - | 2 | 2 (0.8%) |

\*AE=Adverse Events
\*\*SAE=Severe Adverse Events

**Table V.** AE Relationship to the Investigational medicinal product.

|  | At 42 days |  |  |  |  |  | At 120 days |  |  |  |  |  |
| --- | --- | --- | --- | --- | --- | --- | --- | --- | --- | --- | --- | --- |
|  | Vaccine |  |  | Placebo |  |  | Vaccine |  |  | Placebo |  |  |
|  |  | Related/Probably related |  |  | Related/Probably related |  |  | Related/Probably related |  |  | Related/Probably related |  |
| **Adverse Events** | Total | Count | Percent | Total | Count | Percent | Total | Count | Percent | Total | Count | Percent |
| Any AE | 1844 | 1331 | 72.2% | 501 | 321 | 64.0% | 697 | 44 | 6.3% | 268 | 13 | 4.8% |
| **AE\* intensity** |  |  |  |  |  |  |  |  |  |  |  |  |
| Mild | 1664 | 1214 | 72.9% | 449 | 283 | 63.0% | 638 | 37 | 5.8% | 232 | 13 | 5.6% |
| Moderate | 119 | 85 | 71.4% | 28 | 21 | 75.0% | 52 | 7 | 13.4% | 32 | 0 | 0% |
| Severe | 60 | 32 | 53.3% | 20 | 14 | 70.0% | 7 | 0 | 0% | 3 | 0 | 0% |
| potentially life threatening | 1 | 0 | 0.0% | - |  |  | - | - | - | 1 | 0 | 0% |
| death | - | - | - | - |  |  | - | - | - | - | - | - |
| **Any SAE\*\*** |  |  |  |  |  |  | 3 | 0 | 0% | 3 | 0 | 0% |

\*AE=Adverse Events
\*\*SAE=Severe Adverse Events

AEs were further extensively described in terms of events occurrence according to the MedDRA coding system. At the day 42 time point, most of AEs among both the vaccine and placebo groups were general disorders and administration site conditions such as increased temperature and administration site pain. Increased temperature occurred in 47.7% (n=355) of the vaccine group versus 49.6% (n=122) in the placebo arm. Administration site pain reported by 20.9% (n=156) versus 5.4% (n=14), respectively. Vascular disorders such as systolic hypertension occurred in 22.0% (n=164) in the vaccine versus 25.2% (n=62) in the placebo group. Nervous system disorders such as headache occurred in 19.1% (n=142) and 12.2% (n=30), respectively. After 120 days, the main AEs were high temperature (11.7% in vaccine vs. 11.8% in placebo arm) and fever (4.0% vs. 7.7%, respectively). Systolic hypertension occurred in 4.0% vs. 4.5% in the vaccine and placebo groups. Viral infections, such as COVID-19, occurred in 9.3% (n=69) and 11.0% (n=27), respectively. After 180 days, 92 participants tested positive for COVID-19 infection, 62 (8.3%) in the vaccine and 30 (12.2%) in the placebo arms.

Serious AEs (SAEs) were reported only in the placebo arm at 42 days. One subject reported tachypnea as a respiratory disorder and it was related to COVID-19 infection; another subject reported COVID 19 infection, a third subject reported influenza and a fourth had an elevated alanine transferase level (**Table VI**). After 120 days, 3 SAEs were reported in each of the groups. In the placebo arm, a subject reported a lymphoma, another subject reported COVID-19 infection while the third subject reported a hemorrhoidectomy. In the vaccine group, one subject reported back pain, a second reported a maculopapular rash, while a third subject reported chronic cholecystitis (**Table VI**).

**Table VI.**
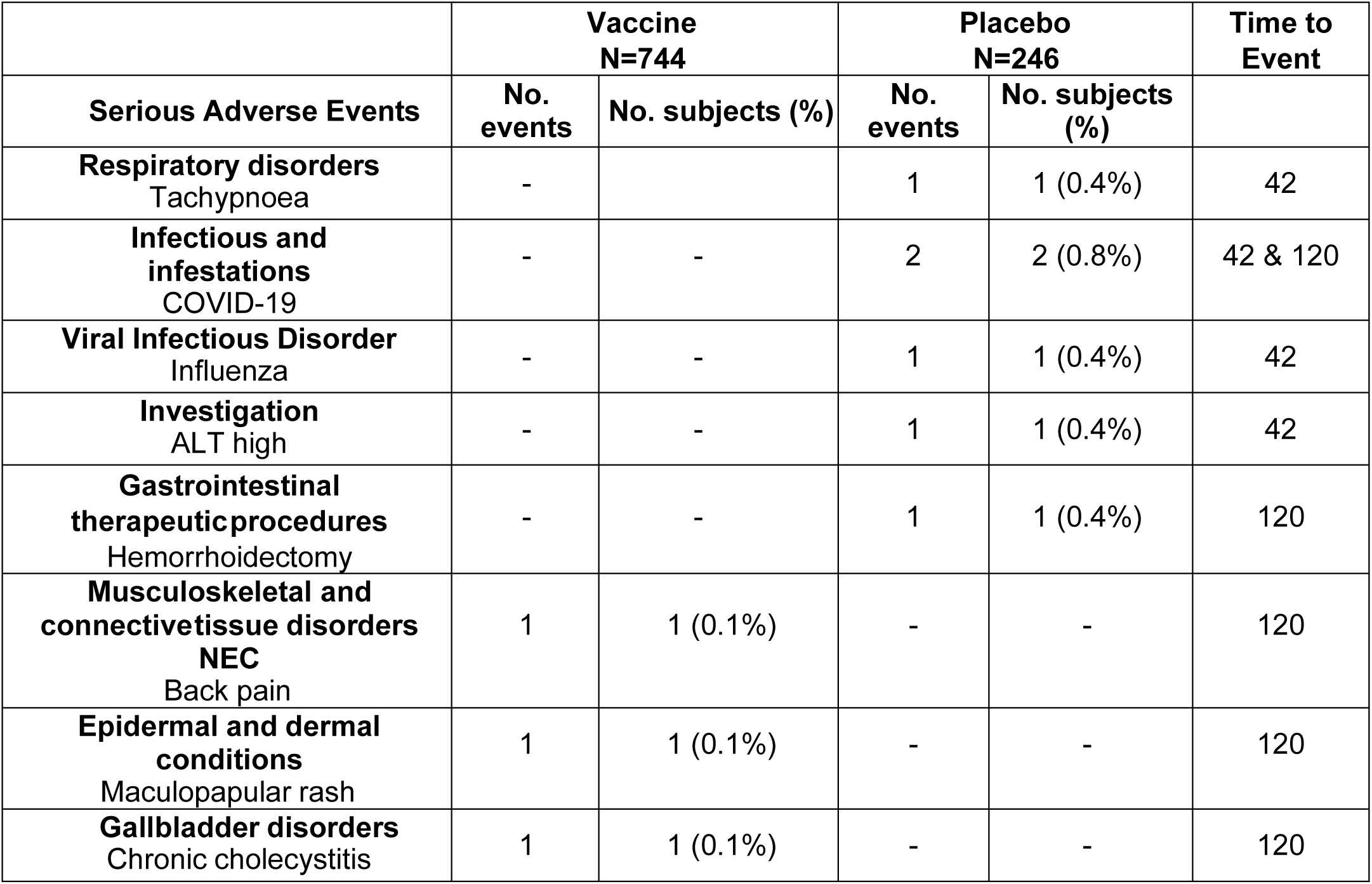
Serious adverse events frequency and percent of participants.

|  | **Vaccine**<br>**N=744** |  | **Placebo**<br>**N=246** |  | **Time to**<br>**Event** |
| --- | --- | --- | --- | --- | --- |
| **Serious Adverse Events** | **No.**<br>**events** | **No. subjects (%)** | **No.**<br>**events** | **No. subjects**<br>**(%)** |  |
| **Respiratory disorders**<br>Tachypnoea | - |  | 1 | 1 (0.4%) | 42 |
| **Infectious and**<br>**infestations**<br>COVID-19 | - | - | 2 | 2 (0.8%) | 42 & 120 |
| **Viral Infectious Disorder**<br>Influenza | - | - | 1 | 1 (0.4%) | 42 |
| **Investigation**<br>ALT high | - | - | 1 | 1 (0.4%) | 42 |
| **Gastrointestinal**<br>**therapeutic procedures**<br>Hemorrhoidectomy | - | - | 1 | 1 (0.4%) | 120 |
| **Musculoskeletal and**<br>**connectivetissue disorders**<br>**NEC**<br>Back pain | 1 | 1 (0.1%) | - | - | 120 |
| **Epidermal and dermal**<br>**conditions**<br>Maculopapular rash | 1 | 1 (0.1%) | - | - | 120 |
| **Gallbladder disorders**<br>Chronic cholecystitis | 1 | 1 (0.1%) | - | - | 120 |

The immunogenicity assessment demonstrated that the vaccine induced an immune response in participants. Sera for detection of SARS-CoV-2 -specific IgG antibodies to S and N Proteins were collected before immunization (on day 1), and at days 42, 120 and 180 after the first dose of vaccine/placebo administration. At baseline, both vaccine and placebo groups had similar levels of IgG antibodies to S protein, with geometric mean (GM) plus 95% confidence interval (CI) of 1.232 (1.155-1.316) for placebo and 1.236 (1.18-1.294) for the vaccine group (p = 0.346). After 42 days, the GM 95%CI was 89.29 (85.46-93.29) for the vaccine group and 1.729 (1.486-2.012) for placebo (p <0.0001). The same trend was also evident at 120 days, with an average of 46.1 for vaccine group and 1.91 for placebo group, and at day 180 showing an average of 30.9 for vaccine group and 2.09 for the placebo group, with the differences being highly significant at both time points (**Figure 2A**). In sharp contrast, IgG antibodies to the viral N protein were not evident in either the vaccine or the placebo group. Both groups had similar antibody concentrations, with GM and 95% confidence interval (CI) of 0.03 (0.029-0.032) for vaccine and 0.03 (0.029-0.038) for the placebo arm (p=0.212). After 42 days, the GM 95%CI was 0.039 (0.036-0.0432) and 0.045 (0.038-0.054) for the vaccine and placebo groups, respectively (p=0.27). This trend was also observed at days 120 and 180 (difference was also nonsignificant at both dates) (**Figure 2B**).

**Figure 2.**
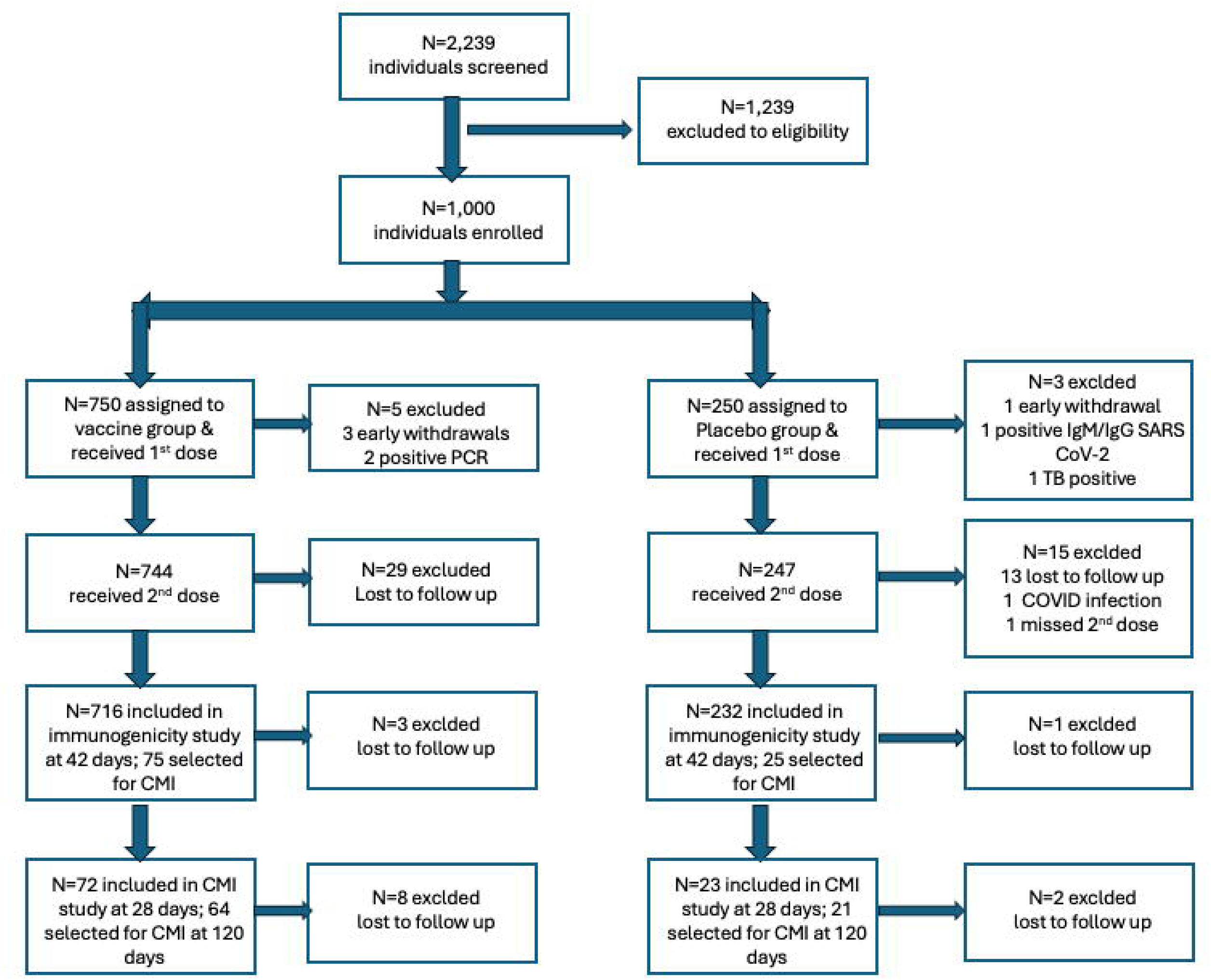
Assessment of the humoral immune response. (**A**) Quantitative IgG to S glycoprotein of SARS-CoV-2 was determined by ELISA (Euroimmun, Lübeck, Germany) on sera collected from participants in the placebo (P) and vaccine (V) study arms on day 1 (before vaccination), and on days 42, 120 and 180 post vaccination. (**B**) Quantitative IgG to N protein of SRAS-CoV-2 was determined by chemiluminescence immunoassay (Abbott, Abbott park, IL) (**C**). Neutralizing antibodies were determined on the same four time points using sera of participants from placebo and vaccine groups. The assay was carried out using general immune parameters (Virus-neutralizing activity (sVNA) (GenScript Biotech, Singapore). The values for individual participants and mean ± SEM are shown in each graph. Asterisks denote statistically significant differences between the indicated groups; **** (p ≤ 0.0001).

Seroconversion, defined as a 4-fold increase in quantitative S protein-specific IgG, was calculated at baseline and in days 42, 120 and 180. Seroconversion was zero at baseline with 95% confidence between (zero and 0.005%) for both arms. After 42 days, the vaccine group had a seroconversion rate (95%CI) of 99.3% (98.4%-99.8%) while the rate in placebo group was 9.4% (6.0%-13.8%), a difference that was highly significant (p<0.001). The unadjusted difference in rate attributed to the vaccine was 90.0%. Similarly, the 4-fold increase was maintained in the vaccine arm at days 120 and 180 with seroconversion rates of 97.1% (95.5%-98.3%) and 94.7% (92.3%-96.5%), respectively. The unadjusted difference in rate versus the placebo was 83.7% at 120 days and 78.9% at 180 days, with significant p-values (p<0.001) (**Figure 3**).

**Figure 3.**
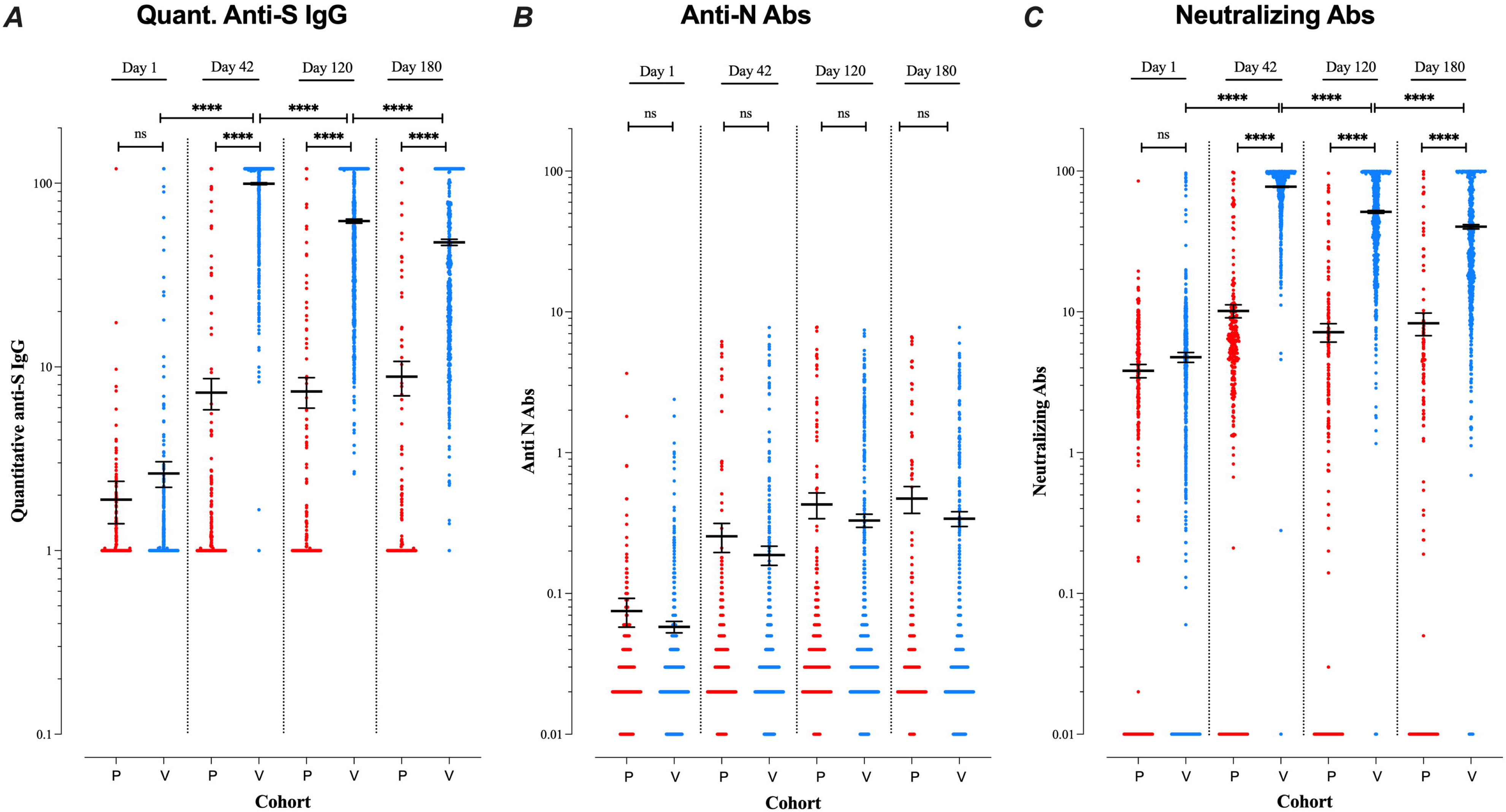
Seroconversion rates in study cohorts. Determination of seroconversion rates was estimated based on a 4-fold increase in the quantitative anti-S protein IgG levels for the placebo and vaccine groups at each of the indicated time points. Asterisks denote statistically significant differences between the indicated groups, *** (p ≤ 0.001), calculated based on Pearson chi^2^.

Neutralizing antibodies to SARS-CoV-2 were determined in vaccine and placebo arms at baseline and at days 42, 120 and 180 post vaccination (**Figure 2C**). Geometric mean titers of neutralizing antibodies showed similar titers at baseline, 0.565 (0.392-0.812) and 0.595 (0.483-0.734) for placebo and vaccine arm, respectively (p=0.186). The levels of neutralizing antibodies increased after immunization, reaching 70.7 (67.49-74.16) (GM with 95%CI) in vaccine group and 3.24 (2.441-4.311) in placebo arm after 42 days (p<0.0001). After 120 days, GM dropped to 38.39 (35.41-41.63) in vaccine group and 0.352 (0.225-0.553) in the placebo arm (p<0.0001). The difference between the two groups was also significant (p<0.0001) at day 180, with a GM of 24.23 (21.36-27.49) for vaccine group and 0.415 (0.245-0.704) for placebo group (**Figure 2C**).

Neutralizing antibodies were also compared between the two study arms of convalescents. A total of 17 subjects (Vaccine group=9 and Placebo group = 8) contracted COVID-19 infection during the study duration until day 42. At baseline, GM 95%CI was 1.99 (1.03-3.89) in the vaccine group and 2.45 (1.08-5.62) in the placebo group. Geometric mean titers of neutralizing antibodies (GM 95%CI) increased after immunization reaching 89.13 (79.43-100) in the vaccine group and 53.70 (36.30-77.62) in the placebo group (p <0.008) at day 42. This indicates the significantly elevated titers of neutralizing antibodies in the vaccinated participants compared to unvaccinated individuals following SARS-CoV-2 exposure.

The cell-mediated response was evaluated in a randomly selected cohort of 98 participants representing the vaccine (n=78) and placebo (n=20) groups on day 1 (before the priming vaccination). The number of participants decreased to 94 participants (vaccine group n=74; placebo n=20) on day 28 (7 days after the booster vaccination) and 84 participants (vaccine group n=66; placebo n=18) on day 120 after the priming vaccination. T cell responses were determined by measuring IFN-γ secretion in cultures of peripheral blood mononuclear cells (PBMCs) after recall stimulation with SARS-CoV-2 S glycoprotein (**Figure 4**). Levels of IFN-γ at baseline were similar for the vaccine group (median 3.04 pg/mL [IQR 2.0-6.15]) and the placebo group (median 2.08 pg/mL [IQR 2.0-6.72]) with a non-significant difference (p=0.241). After 28 days, PBMCs of participants in the vaccine group secreted substantially higher levels of IFN-γ after restimulation (median 94.01 pg/mL [IQR 50.28-211.6]), which was significantly different (p=0.003) in comparison with the placebo group (median 10.55 pg/mL [IQR 2.69-26.65]). The day 28 response of the vaccine group was also significantly different (p <0.0001) compared to that on day 1, the day of administration of the first dose (**Figure 4**). Importantly, T cell recall responses were still evident after 120 days in the vaccine group (median 61.87 pg/mL [IQR 12.85-210.5]) compared to the placebo group (median 12.94 pg/mL [IQR 4.83-50.56]) (p<0.039). The day 120 response of the vaccine group was still significantly different (p <0.0001) compared to the day 1 response but was not significantly different from the day 28 response (p=0.114).

**Figure 4.**
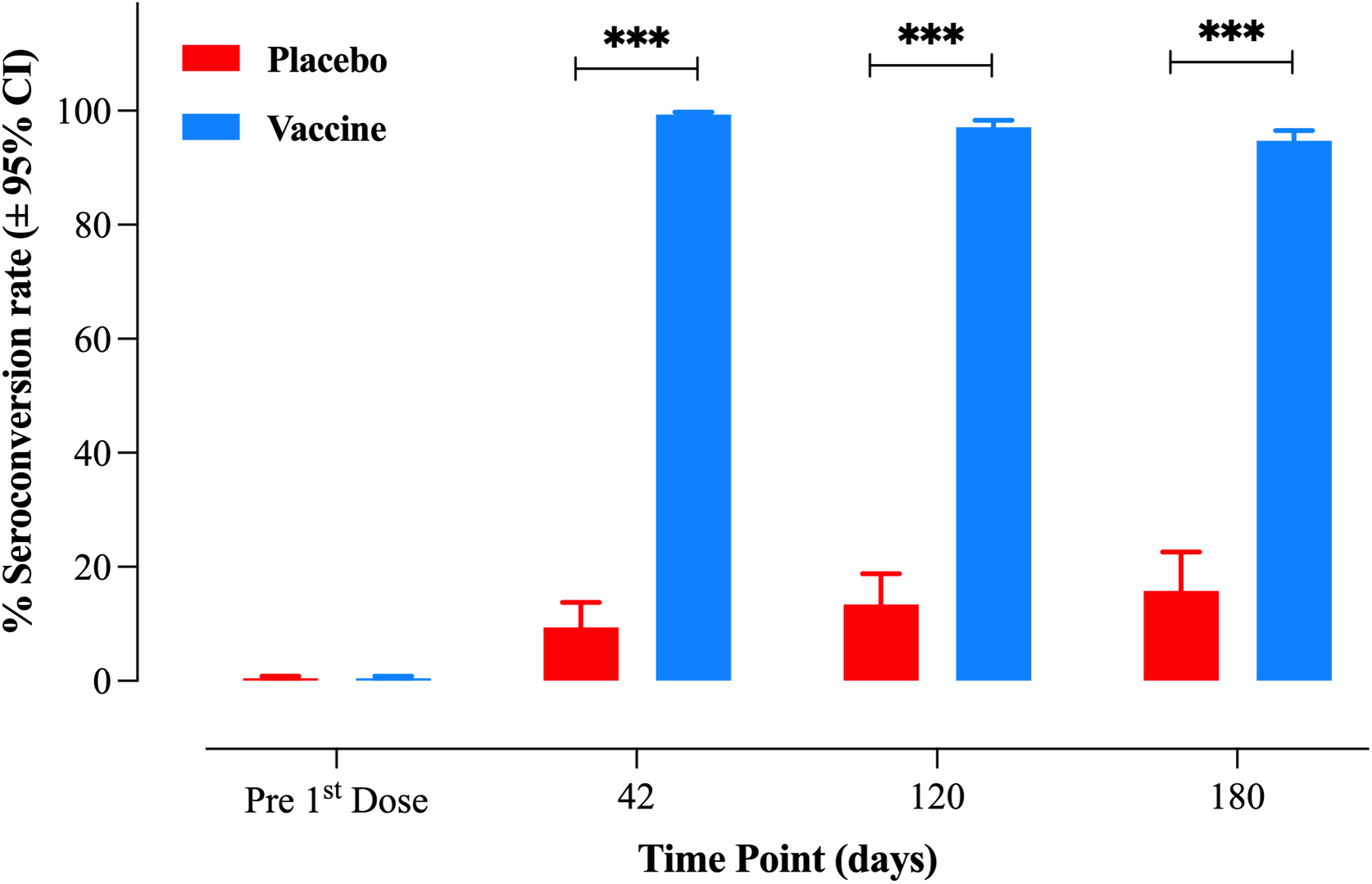
Assessment of cell-mediated IFN-γ response to SARS-CoV-2. The extent of IFN-γ production in supernatants collected after culturing peripheral blood mononuclear cells in the absence (control) or presence of SARS-CoV-2 S glycoprotein (SP) for 72 hr. PBMC were collected from participants of both study arms on days 1, 28 and 120 post vaccination. The amount of IFN-γ in culture supernatants was determined using a human IFN-γ-specific ELISA (Cat# KAP1231 from DIAsource ImmunoAssays), as per the recommendation of the manufacturer. The values for individual participants and mean ± SEM are shown. Asterisks denote statistically significant differences between the indicated groups; * (p ≤ 0.05), ** (p ≤ 0.01), and **** (p ≤ 0.0001).

In order to assess the level of cell mediated immune responses in a T cell type-specific manner, we measured the extent of cell proliferation in response to antigen-stimulation in both T-helper (CD4^+^) and T-cytotoxic (CD8^+^) cells by flow cytometry. Cellular immune responses showed proliferation of antigen-specific cells of both CD4^+^ and CD8^+^ T cells in response to glycoprotein S, particularly on day 28 (**Figure 5**). The CD4^+^ T cell responses of the placebo and vaccine groups at day 1 were at baseline with no significant difference between them (p=0.433). The CD4^+^ T helper response at day 28 reached a median proliferation of 0.14% [IQR 0.01-0.325] in the vaccine group compared to a median of 0.02% [IQR 0.01-0.07]) in the placebo group (p=0.0014) (**Figure 5A-B**). Similarly, there was a significant enhancement in the antigen-driven proliferation of CD4^+^ T cells on day 28 compared to day 1 of the vaccine group, the latter having a median of 0.035% [IQR 0.01-0.182] (p=0.007). Interestingly, the CD4^+^ T cell proliferative response weakened considerably by day 120, reaching a median of 0.09% [IQR 0.01-0.285], that had no statistical significance compared to day 1 responses (p=0.097) (**Figure 5B**). In contrast, the CD8^+^ T cell proliferative response in vaccinated individuals was significantly elevated on day 28 (median 0.1% [IQR 0.01-0.23]) (p=0.022) and day 120 (median 0.11% [IQR 0.01-0.3]) (p=0.001) compared to day 1 (**Figure 5C-D**), suggesting that the S protein-specific CD8 T cell response is more durable following vaccination than the CD4 T cell response. One noticeable finding in the placebo participants was the apparent rise in CD8 T cell response observed at day 120 (median 0.085 % [IQR 0.01-0.255]) compared to day 1 (p=0.038). This is most likely due to natural exposure to SARS-CoV-2 virus in the community. In fact, at least 2 of the 20 participants in the placebo group had evidence of positive PCR tests for the virus between days 28 and 120 of the trial.

**Figure 5.**
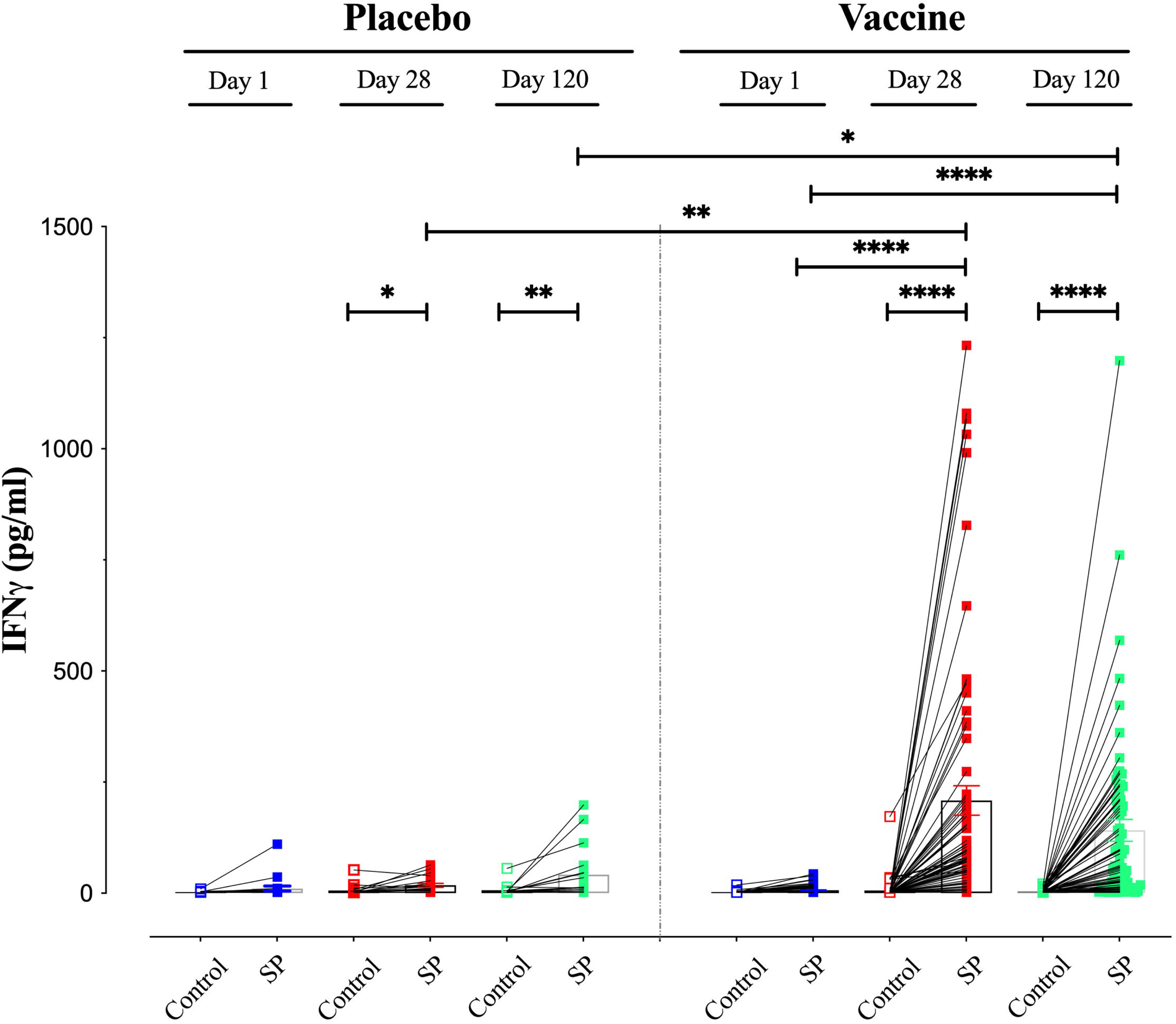
Assessment of CD4^+^ and CD8^+^ T cell proliferative responses to SARS-CoV-2 S protein. Cellular proliferation was determined following incubation of PBMCs for 72 hr with or without the viral S protein using CFSE labeled cells. At the end of culture period, cells were harvested, stained with monoclonal antibodies specific to CD4 or CD8 protein (Biolegend) and analyzed by flow cytometry. Proliferating (with reduced CFSE staining) CD4^+^ and CD8^+^ Т lymphocytes were detected in the cell mixture using a BD FACSCelesta flow cytometer. The values for individual participants and mean ± SEM are shown. Asterisks denote statistically significant differences between the indicated groups; * (p ≤ 0.05), and ** (p ≤ 0.01).

## DISCUSSION

In this study, we report the findings on the safety and immunogenicity of Gam-COVID-Vac evaluation from analysis of this phase II/III clinical trial conducted in the UAE. The vaccine efficacy was consistent with previous studies of Sputnik-V vaccine in other populations (8, 13)(14). Our findings show that the vaccine was well tolerated, with only 0.4% of participants in the vaccine group reporting serious adverse events, all of which were considered to be unrelated to the vaccine. No deaths were reported. The data show that revaccination may cause milder adverse events compared to first dose of immunization against COVID-19. The AEs observed after the two doses and after 120 days of the study vaccine are typical reactions to most types of vaccines, with the great majority being in the mild/moderate categories, including general disorders and administration site conditions. The COVID-19 case incidence among the vaccine and placebo groups was 62 (8.3%) versus 30 (12.1%), respectively, showing a partially protective effect after a two-dose immunization; however, the study design does not allow us to draw conclusions from these observations. Of note, the observed adverse effects using the Sputnik V vaccine are milder than those observed using the ChAdOx1-S nCoV-19 or mRNA vaccines. According to published reports, serious thrombosis with thrombocytopenia syndrome (TTS) mediated by platelet-activating antibodies against platelet factor 4 (PF4), which clinically mimics autoimmune heparin-induced thrombocytopenia, were reported following the ChAdOx1-S nCoV-19 vaccine (15). Those symptoms were similar to those observed in patients developing Long COVID following infection. Moreover, recent evidence has suggested that mRNA vaccines may induce persistent, and, in some cases, severe adverse effects in a small percentage of individuals (16). No symptoms of long COVID were reported by the nearly 1000 participants in our study.

The immunogenicity of the Gam-COVID-Vac vaccine was demonstrated by the induction of strong humoral and cellular immune responses (8). The level of virus neutralizing antibodies is thought to be a predictive indicator of immune protection against COVID-19 but the protective cutoff value remains unknown (17). In the current report, the neutralizing antibody response was monitored at days 42, 120, and 180 after immunization, and the results demonstrate that the sputnik V vaccine effectively induced a strong and specific humoral immune response. Two injections of the Gam-COVID-Vac vaccine induce VNA in 100% of participants by day 42 after immunization. The results indicated that GMR increased and reached maximum at day 120 with an average of 24.14, with a significant difference between the two study arms, followed by a decline at 180 days. Nevertheless, the optimal interval between injections and times of booster injections remains undetermined.

A comparison of the convalescent serum samples after COVID-19 infection in both study arms at day 42 suggests that the observed humoral immune response was likely boosted by vaccination, with GMR 95%CI increasing to 89.13 in the vaccine group compared to 53.70 in the placebo group (p-value<0.008). These findings indicate that the Gam-COVID-Vac vaccine is a good candidate for boosting immune responses (after initial immunization or natural infection) against COVID-19 used in the re-vaccination regimen.

There were notable significance differences in the vaccine effects on immunogenicity. On day 42 after immunization, the vaccine induced an increase in quantitative IgG concentration in 99.3% of participants. This increase was maintained in the vaccine arm at 120 and 180 days with seroconversion rates of 97.1% and 94.7%, respectively. After 42 days, the GMR 95%CI was 54.95 for vaccine group and 1.70 for placebo group, a difference that was highly significant, and this pattern was still evident at 120 and 180 days.

Our findings demonstrate that a 2-dose immunization regimen with the Gam-COVID-Vac vaccine resulted in the formation of anti-viral cell-mediated immune responses, as evidenced by the significant rise in the median of proliferating antigen-specific CD4^+^ and CD8^+^ T-lymphocytes and extent of secreted IFN-γ after vaccination as compared to baseline levels. The significant rise in the percentage of proliferating antigen-specific CD4^+^ T cells was observed on day 28 following the primary vaccine dose, and 7 days after the booster dose. This response appeared to decline by day 120 to a non-significant level but remained marginally higher than baseline. In contrast, there was a significant and persistent rise in antigen-specific CD8 T cell proliferation on day 28 which remained significant on day 120.

Recently, a study compared the neutralizing activity of sera collected from participants after receiving 2 doses of the Gam-COVID-Vac vaccine (n=31) or the BNT162b2 vaccine (n=17) against the omicron variant of SARS-CoV-2 (18). Serum samples were collected longitudinally from 1-2 weeks up to 6 months after the second vaccine dose. The findings demonstrated that the titer of neutralizing antibodies in sera of individuals after Gam-COVID-Vac vaccine persisted longer than in sera of individuals given the mRNA vaccine. In general, the relative efficacy of sera following the Gam-COVID-Vac vaccine was ∼3-fold higher than equivalent sera from mRNA vaccine group. Thus, heterologous vaccination using the Gam-COVID-Vac vaccine could be an effective strategy to boost immunity against SARS-CoV-2 variants in individuals who had received other forms of vaccination, including the BNT162b2 vaccine (18).

While Gam-COVID-Vac had an efficacy of 91.6% (85.6-95.2) to prevent severe COVID-19 after the second dose (8), the single dose Ad26.COV2.S vaccine prevented 66.1% (55.0 to 74.8) of moderate to severe COVID-19 at 28 days (19), and 2 doses of the AZD1222 (ChAdOx1 nCoV-19) vaccine had 74.0% efficacy in preventing COVID-19 (65.3-80.5) (20). In a multi-center trial, the AZD1222 vaccine had an efficacy of 66.7% (57.4% to 74%) against SARS-CoV-2 (21) and prevented 70.4% (43.6 to 84.5) of symptomatic alpha variant COVID-19 in the UK (22). Convidecia™Ad5-nCoV prevented 65.7% of symptomatic COVID-19 28 days after a single dose and was 91% effective in treating severe disease (23).

Our findings concerning AEs are in line with what has been recently reported (24). The reported AEs varied based on the type adenoviral-vectored vaccines. The percentage of AEs reported for Gam-COVID-Vac single dose (Sputnik Light), and Gam-COVID-Vac double dose Sputnik V was estimated to be 85.3% and 82.6%, respectively. Percentage AEs reported for the AZD1222 (AstraZeneca) vaccine tended to be higher (93.9%) and were associated with systemic manifestations, including fever and headache which was reported in more than 50% of participants after receiving the vaccine (21, 24).

In conclusion, the results of this randomized, controlled, phase II/III trial of Gam-COVID-Vac in UAE demonstrate the high immunogenicity and good tolerability profile of the vaccine in participants aged 18 years or older. Our study provides evidence for significant, long-term, cell-mediated immune responses to the vaccine, in particular CD8^+^ T lymphocytes and IFN-γ production, persisting for at least 120 days.

## Limitations

Limitations of this study include being a single-center study with a relatively small sample size compared to other international studies of phase III trials. However, this study was focused on the efficacy of the Gam-COVID-Vac vaccine in the United Arab Emirates which has a unique population of diverse ethnic and genetic backgrounds. Although the number of participants in the study is small, the efficacy of the Gam-COVID-Vac in our cohort is in line with the interim analysis of Phase III conducted on 22,000 volunteers in Russia (8). Another limitation of our study is the lack of information on the neutralization of vaccine-induced sera against other SARS-CoV-2 variants. There is evidence that neutralization capacity of sera following vaccination with ancestral S protein-containing vaccines is reduced when tested against many viral variants (25–27). Nevertheless, when compared with mRNA vaccination, the relative efficacy of Gam-COVID-Vac-induced sera was ∼3-fold higher than equivalent sera from the mRNA vaccine group (18). It is generally believed, however, that T cell immunity against viral variants is maintained, hence forming the basis of continued protection against infection in humans (28, 29). Further studies of candidate vaccines are needed to obtain information on the defining parameters of protection and duration of post-vaccine protective immune responses.

## Author Contributions

Ahmed AK. Al Hammadi: Principal Investigator. Protocol revision, approval, supervision of study, data analysis, manuscript writing and final review, submission.

Amna H. Alzaabi: Sub-investigator. Subject enrolment, review and approval of final manuscript

Haneen B. Choker: Sub-investigator. Subject enrolment, review and approval of final manuscript

Ahmed A. Ibrahim: Sub-investigator. Subject enrolment, review and approval of final manuscript

Ahmed I. Mahboub: Sub-investigator. Subject enrolment, review and approval of final manuscript

Reem S. Al Dhaheri: Sub-investigator. Subject enrolment, review and approval of final manuscript

Mohamed N. Alzaabi: Sub-investigator. Subject enrolment, review and approval of final manuscript

Timothy A. Collyns: Conduction of baseline labs, management of samples and supervision of sample handling and transportation to collaborating central labs; review of final manuscript.

Gehad ElGhazali: Conduction of humoral immune testing and review of final manuscript.

Stefan Weber: Conduction of humoral immune testing and review of final manuscript.

Basel al-Ramadi: Co-Principal investigator; conduction of cellular immune testing, data collection and analysis, manuscript writing and review of final manuscript.

## Data Availability

All data produced in the present work are contained in the manuscript

## Acknowledgments

We wish to thank our global sponsor (Human Vaccine LLC) and the local sponsor in the UAE (Aurugulf Health Investment). We also wish to thank the Russian Ministry of Healthcare FGBU N.F. Gamaleya Scientific Research Center of Epidemiology and Microbiology for all help received to initiate the study and providing logistical support. We wish to thank the team of Infectious Disease department at Tawam Hospital for their support during the study. We are grateful to Dr Shoja Haneefa, Yassir Mohamed, Ashraf Al-Sbiei, Heba Taji and Ienas Idriss (Department of Medical Microbiology & Immunology, College of Medicine & Health Sciences, United Arab Emirates University) for performing the cell mediated immunity assays. We also thank the laboratory staff at Purelab, Sheikh Khalifa Medical City-Abu Dhabi, for performing all of the serological assays. We would like to thank the study participants, clinical site staff, and PDC clinical research organization for their collaboration.

